# The seasonality of varicella in the tropical climates of Colombia: A statistical and mathematical modeling study

**DOI:** 10.1101/2022.12.06.22283152

**Authors:** Laura Andrea Barrero Guevara, Elizabeth Goult, Dayanne Rodriguez, Luis Jorge Hernandez, Benedikt Kaufer, Tobias Kurth, Matthieu Domenech de Cellès

## Abstract

**Background:** Varicella remains a major burden in many tropical regions, where low- to middle-income countries report the highest mortality rates. Understanding when and where varicella incidence increases could help us understand viral transmission and optimize the allocation of resources. Due to the lack of surveillance data, the epidemiology of varicella in the tropics has not been well characterized. Here, we assessed varicella seasonality and analyzed its correlation with climatic variables across Colombia.

**Methods:** We compiled an extensive dataset of weekly varicella reports in children up to the age of ten in 25 municipalities across Colombia. We used generalized additive models to describe the seasonality of varicella in each municipality. Using clustering methods and matrix correlation tests, we then compared the spatial variability in varicella seasonality with that in five meteorological variables across the municipalities. Finally, we developed a mathematical model to examine whether the influence of the climate on transmission rates could result in the observed seasonal patterns of varicella in Colombia and other Central American countries.

**Findings:** Varicella seasonality was markedly bimodal, with a more pronounced peak early in the year in northern municipalities (≈week 15), while later in the year (≈week 40) in southern municipalities, including Bogotá. This spatial gradient was strongly correlated with humidity (Mantel statistic = 0·412, p-value = 0·001) but not temperature (Mantel statistic = 0·077 and p-value = 0·225). Furthermore, a mathematical model that included a small, negative association between humidity and transmission was able to reproduce the observed spatial patterns in Colombia and México. This model also predicted a latitudinal gradient in other tropical countries of Central America, where the seasonality of varicella has not yet been characterized.

**Interpretation:** These results demonstrate a large variability in varicella seasonality across the tropical climates of Colombia. They further suggest that seasonal fluctuations of humidity explain the varicella epidemics calendar in Colombia and other Central American countries. More generally, our results highlight the need to carefully consider the subnational heterogeneity of climates when studying the seasonal epidemiology of varicella and assessing the impact of control measures.

**Funding:** Core funding from the Max Planck Institute for Infection Biology of the Max Planck Society, Berlin, Germany.

## Introduction

The varicella-zoster virus (VZV) causes varicella (chickenpox), a disease that afflicts over 70 million children every year worldwide.^1^ Although the disease is typically mild and self-limiting, it can result in pneumonia, encephalitis, sepsis, and death (over 14,000 estimated deaths worldwide in 2019),^1^ particularly in immunocompromised children.^2^ After primary infection with VZV, the virus establishes latency in sensory neurons. Later in life, the virus can reactivate and cause shingles, a debilitating painful disease particularly prevalent in the elderly (2·9-19·5 cases per 1,000 individuals over 50 years old).^3^ Hence, understanding the epidemiology of varicella, not just because of its burden but also that of shingles, highlights the need to better characterize the epidemiology of varicella.

As for many other infectious diseases, the incidence of varicella is highly seasonal, with a latitudinal gradient in the peak timing observed on a global scale.^4^ In temperate regions, this seasonality is typically characterized by a single peak at the end of winter (e.g. in the US and Germany),^5,6^ although two peaks have also been described (e.g. in Japan and China).^7,8^ The seasonal patterns in subtropical regions generally mirror those in temperate regions.^9^ In tropical regions, however, varicella seasonality appears to be less definite, although observations have remained scarce because of a lack of epidemiological surveillance.^10^ Bridging this data gap in tropical regions is crucial for two reasons. First, most of the morbidity and mortality associated with varicella affect low- to middle-income countries, many of which have a tropical climate.^1^ Second, the climate in tropical regions—with almost constant temperature but variable precipitation and humidity during dry and rainy seasons—offers an opportunity to evaluate how different climatic variables impact varicella transmission, expanding on previous studies in temperate regions.^7,11^ A better understanding of varicella seasonality in a given setting may support public health efforts, informing epidemic preparedness and infection control.

In this study, we aimed to characterize the calendar of varicella epidemics across the climatically diverse regions of Colombia, where exhaustive surveillance has been in place for several years. We then used statistical models to assess whether climatic variables were correlated with varicella seasonality. Finally, we examined whether these correlations could be explained by mathematical models with climatically-forced seasonality in varicella transmission.

## Methods

### Data sources

#### Varicella data

We accessed the clinically confirmed varicella notifications freely available from the Colombian national surveillance system (*Sistema Nacional de Vigilancia en Salud Pública*, SIVIGILA, for more details, please refer to the supplementary data). By focusing on the 2011–2014 pre-vaccination period, we reduced the reporting bias from the SIVIGILA implementation in 2007.^12^ Additionally, we focused on children up to the age of ten to avoid zoster cases and changes in seasonality as a consequence of children moving into secondary school. As both age and birthdates are recorded in the system, we only considered cases for which the difference between the age reported and the age calculated from the date of birth did not exceed one year.

Colombia is divided into 1122 municipalities, administrate divisions roughly equivalent to counties. As many municipalities had low case counts, for definiteness, we selected the municipalities with a signal-to-noise ratio (mean to standard deviation ratio of the weekly reports) above one and an average of at least five varicella cases/week. 25 municipalities met these criteria (Fig. S1), which collectively covered 67·4% of the total reports of varicella (range 64·6% in 2013 to 69·7% in 2011) and 41·2% of the total Colombian population up to ten years old during the study period (range 41·0% in 2014 to 41·3% in 2011). These 25 municipalities included a wide range of climates (latitude range 1·20–11·00° N, longitude range 72·5–77·3° W, and six of the 17 Köppen–Geiger climate classifications most commonly found in the tropics) and had an average area of 686·8 km^2^ (range 17 km^2^ in Itagüí to 3,141 km^2^ in Montería (Fig. S1)).

#### Demographic data

We obtained the weekly varicella incidence among children up to ten years per municipality using the annual demographic estimates from the Colombian National Administrative Department of Statistics (DANE).^13^

#### Climate data

Weekly mean temperature (°C), specific humidity (defined as the mass of moisture per mass of air), and relative humidity (%) data were extracted from the 32 km grid North American Regional Reanalysis (NARR) dataset, and the total weekly precipitation from the 10 km grid Climate Hazards Group InfraRed Precipitation with Station (CHIRPS) dataset for each municipality. In addition, we calculated the absolute humidity (defined as the mass of water per volume of air) from the extracted relative humidity (Fig. S2) considering the altitude of the municipalities.^14–16^ For Mexico and Central America countries (Panamá, Costa Rica, Nicaragua, Honduras, El Salvador, Guatemala, and Belize), we obtained the specific humidity from the NARR for the capital city of every first-level subnational division.

### Descriptive model of varicella seasonality

To describe the spatial variability in varicella seasonality across Colombia, we used generalized additive models (GAMs). These flexible extensions of generalized linear models allow the modeling of complex, potentially non-linear associations using smooth functions of predictor variables while preventing overfitting by penalizing the wiggliness (quantified by the second derivative) of the function.^17^ The base model included two predictors: a cyclic spline term (with a period of 1 year) to model the seasonality in each municipality and a municipality-year parametric intercept to capture the long-term trends in each municipality. The model included a population size offset (log(population)), effectively modeling the incidence rate. To model the spatial variability in varicella seasonality, we included a tensor product smooth between the week number and latitude with two smoothing bases: a cyclic spline for the week number and a cubic regression spline for the latitude. For all models, we used maximum likelihood (ML) to estimate the parameters of all models,^18^ and we calculated the Akaike information criterion (AIC) to compare their parsimony.

### Correlation between varicella seasonality and climatic variables

We aimed to assess the correlation between the spatial variability of varicella seasonality and that of climates across Colombia. We did not attempt, however, to directly regress against meteorological variables because seasonal forcing in transmission and long-term changes in population immunity can result in complex, non-linear incidence dynamics that may not be easily captured by standard regression models.^19^ Instead, we first calculated the similarity matrix (where similarity was quantified using the Euclidean distance) of varicella data and of each climate variable between municipalities across Colombia.

We used the similarity matrices to cluster the municipalities and evaluate the correlation between the climatic variables and varicella. We clustered each dissimilarity matrix using the partitioning around medoids (PAM) algorithm. We used the visual assessment of cluster tendency (VAT) and Hopkins statistics to inspect the clustering tendency of the data and defined the number of clusters using the average silhouette mean and the elbow method. Finally, we used Mantel tests to estimate the correlation between each pair of dissimilarity matrices.

### Transmission models of varicella seasonality

To further test and formalize the hypothesis that climatic variables explain the seasonality of varicella in Colombia, we developed a simple compartmental transmission model (SEIR model, for more details, please refer to the supplementary methods), in which the transmission rate of varicella infection was seasonally forced by climate and by changes in contact rates due to alternating school terms and school vacations. The model was represented as a set of differential equations describing the movement of individuals between the compartments. Briefly, individuals started at the susceptible to varicella compartment (S). Next, susceptible individuals entered the exposed compartment E with a seasonal force of infection λ (per susceptible rate of infection, reproduction number, 10).^20^ Then, exposed individuals become infectious at a latency rate (1/σ = average incubation period, 1/14 days),^21^ enter the infectious compartment I, and then move to the R recovered compartment at a rate (1/γ = average recovery period, 1/7 days).^21^ Furthermore, we used the same model together with data from climatic variables from México and other countries in Central America to explore the seasonal varicella transmission in these subtropical and tropical countries.

### Numerical implementation

R version 4.1.2 (2021-11-01) was used for all analyses. The climate data were obtained using the packages “ncdf4”, “chirsp”, “humidity”, and “kgc”.^15,22–24^ Colombia map figures were created using the package “colmaps”.^25^ The package “mgcv” was used for all the GAMs estimations.^17^ The dissimilarity matrices, clusters, and mantel tests were obtained with the packages “TSclust” and “vegan”.^26,27^ The simulations were performed using the “pomp” package.^28^ All data are freely available on the web pages of the SIVIGILA, IDEAM, NARR, and CHIRPS. Furthermore, all the aggregated data and code are stored in Edmond to ensure the reproducibility of the results.

## Results

### Incidence of varicella in Colombia between 2011 and 2014

Between 2011 and 2014, a total of 421,085 varicella cases were reported in Colombia. First, we eliminated cases with missing age information or a mismatch between age and birthdate (0·018% and 0·097%, respectively, Fig. S3). Then, we selected only the cases from children up to ten years, representing 55·5% of the cases throughout the country. After applying the municipality selection criteria, the final dataset comprised 156,976 cases in 25 municipalities (67·4% from the cases in children up to age 10), which spanned a large area of Colombia (latitude range: 1·20–11·00° N, longitude range: 72·49–77·28° W, Fig. 1A and S1).

**Figure 1.**
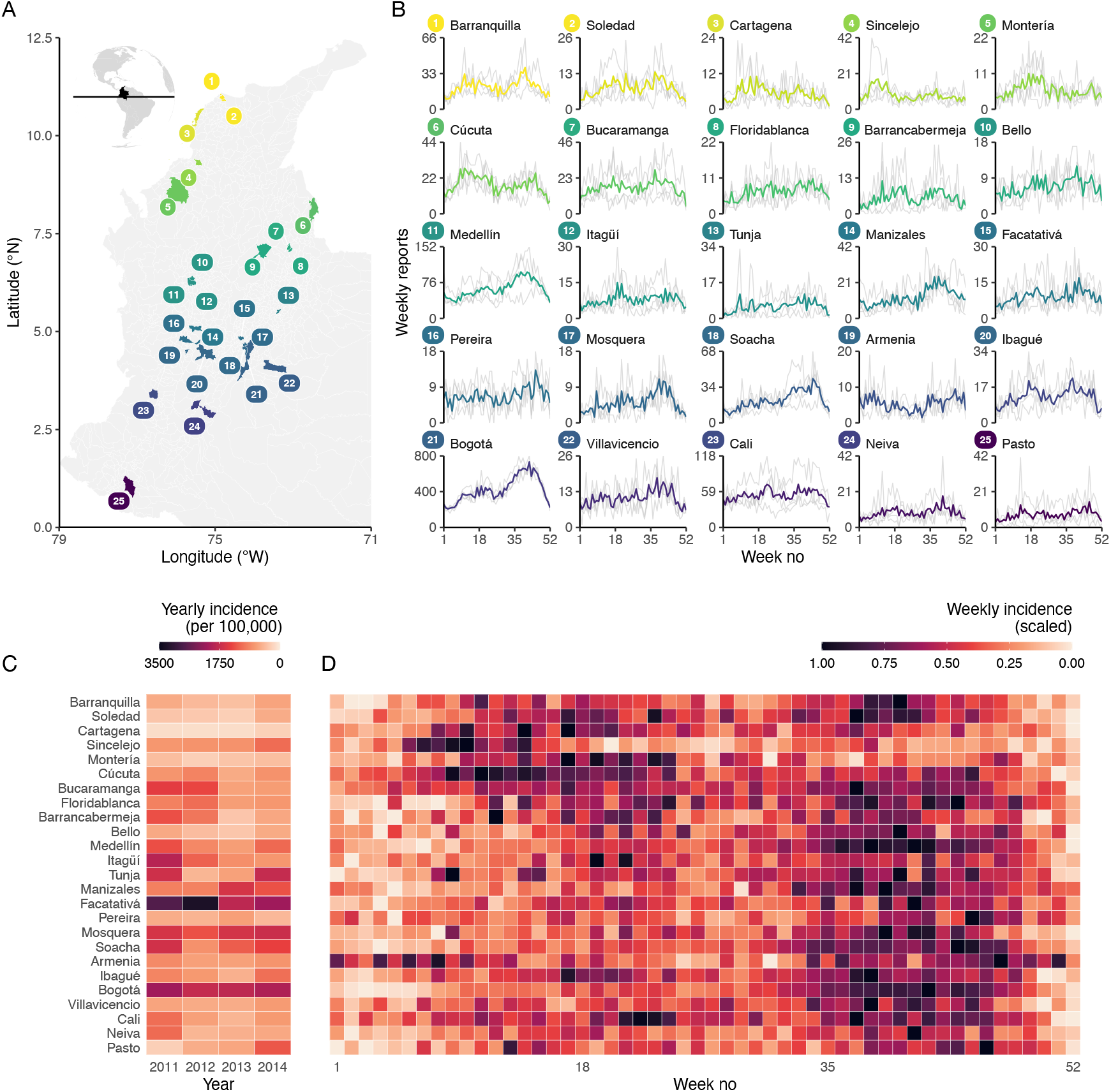
Varicella reports and incidence of varicella in Colombia, 2011–2014. (A) Map of included municipalities. (B) Mean weekly varicella reports in the municipalities of Colombia, in gray are the weekly reports every year. (C) Mean yearly varicella incidence per 100,000 children up to ten years old per municipality, and (D) Mean weekly varicella incidence of varicella per municipality (rescaled so that 0 is the minimum and 1 is the maximum number of cases per municipality). From bottom to top, the municipalities are ordered by increasing latitude.

Most varicella cases were reported in Bogotá (56·1% of the total included cases), followed by Cali (8·30%) and Medellín (7·06%), the three most populated municipalities in Colombia (Fig. 1B). The yearly reported incidence varied substantially (range, 3291·8 cases per 100,000 children per year in Facatativa in 2012 to 113·9 cases per 100,000 children per year in Cartagena in 2012) (Fig. 1C). However, the mean age of infection was similar across municipalities, ranging from 4·6 to 5·5 years (Fig. S3).

### Latitudinal gradient demonstrates substantial spatial heterogeneity of varicella seasonality across Colombia

The incidence of varicella was markedly seasonal across Colombia, with an early peak around week 15 and a late peak around week 40. However, the amplitude of both peaks changed substantially with latitude (Fig. 1D). The best-fitting GAM confirmed the spatial variability in varicella seasonality, with strong statistical evidence of a latitudinal gradient (Fig. 2A and S4, ΔAIC = 309 compared with a model with no spatial variability). The early peak was more pronounced than the late peak in northern (latitude range: 7·00–11·00° N) municipalities, for example, in Cúcuta (7·88° N latitude, Fig. 2B and C). By contrast, the late peak had higher amplitude in southern (latitude range 1·20–7·00° N) municipalities, such as Bogotá (4·71° N latitude, Fig. 2B and C). The best-fitting model correctly reproduced the observed data in every municipality (overall R-squared: 71·9% in Soacha to 98·5% in Mosquera). Hence, our descriptive model accurately recapitulated the spatiotemporal dynamics of varicella and evidenced substantial spatial heterogeneity of varicella seasonality across Colombia.

**Figure 2.**
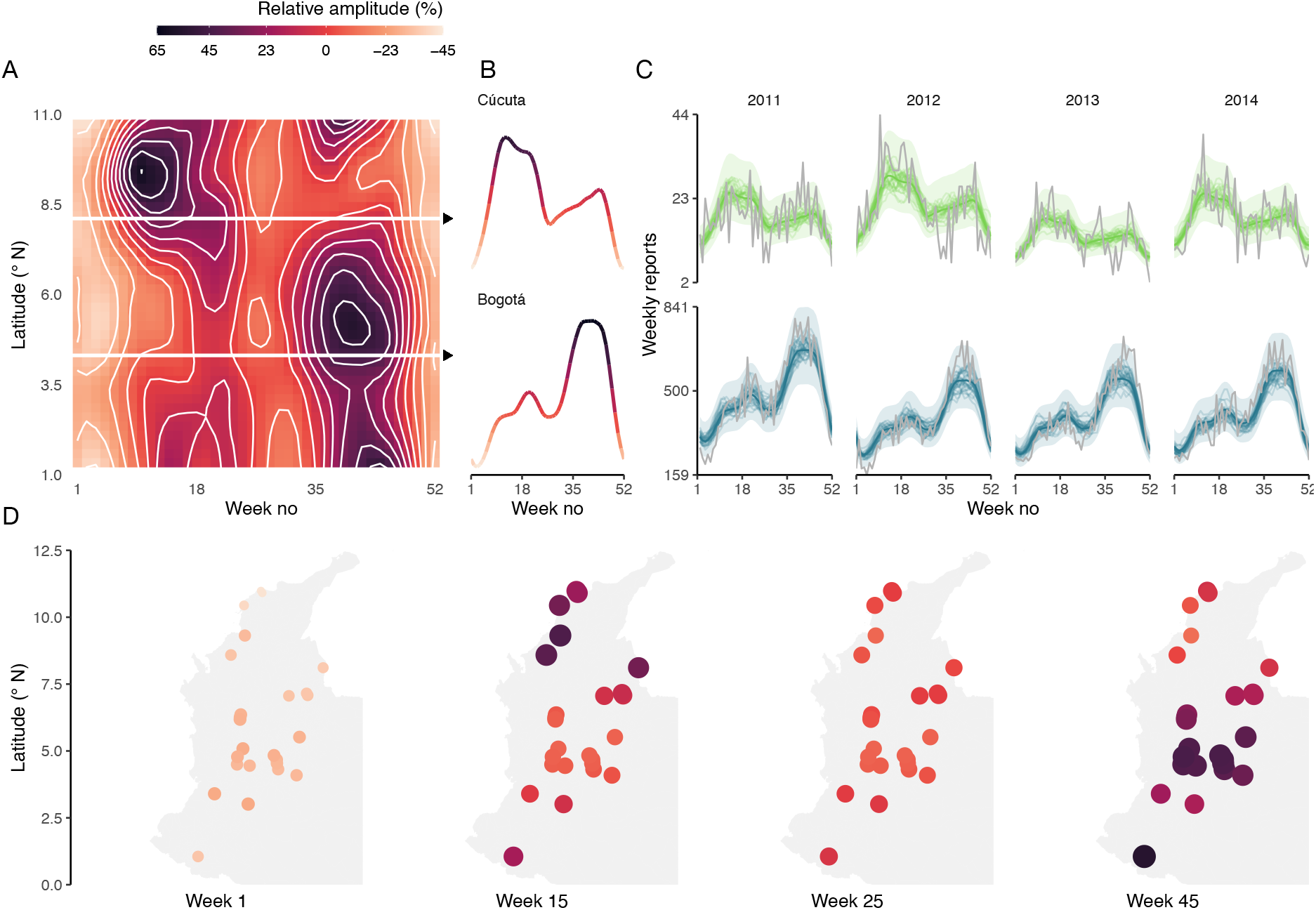
Estimated latitudinal gradient of varicella seasonality across Colombia. (A) The shape of the partial effects from the seasonality (relative amplitude (%) from the mean) for varicella incidence across latitudes. (B) Estimated seasonality in Cúcuta (7·88° N latitude) and in Bogotá (4·71° N latitude). (C) Observed (gray lines) and predicted (colored lines and envelopes) seasonality of varicella reports in Bogotá and Cúcuta throughout the study period. The lines represent 30 random draws from the model posterior, and the ribbons represent an approximate 95% simultaneous confidence interval for the fitted GAM. (D) Spatial variability in the varicella seasonality (relative amplitude) across municipalities at the beginning (week 1) and middle of the year (week 25) and at the varicella peaks (weeks 14 and 45).

### Spatial variability of the varicella seasonality correlates with variability in climates across Colombia

The latitudinal gradient described above suggests that spatial heterogeneity of climates could explain the observed seasonality of varicella across Colombia. We formalized this hypothesis of a causal framework using a directed acyclic graph (DAG), in which latitude explained climate, which itself explained varicella seasonality (Fig. S6). To test this hypothesis, we first applied clustering methods to identify groups of municipalities with broadly similar varicella seasonality. In keeping with our previous observations, we found evidence for two clusters (Hopkins statistic = 0·69), one including northern municipalities with a more pronounced early peak and the second including southern municipalities with a more pronounced late peak (Fig. 3B). Next, we clustered the data for each climate variable (Fig. 3B). For specific humidity, absolute humidity, and precipitation, we found evidence for two clusters (Hopkins statistic = 0·75, 0·74 and 0·79 respectively), which broadly matched the two clusters of varicella seasonality (fraction of matching pairs: 81·8%, 81·8%, and 95·5% respectively). By contrast, the temperature and relative humidity variables clustered into more than two groups that did not match those of varicella seasonality.

**Figure 3.**
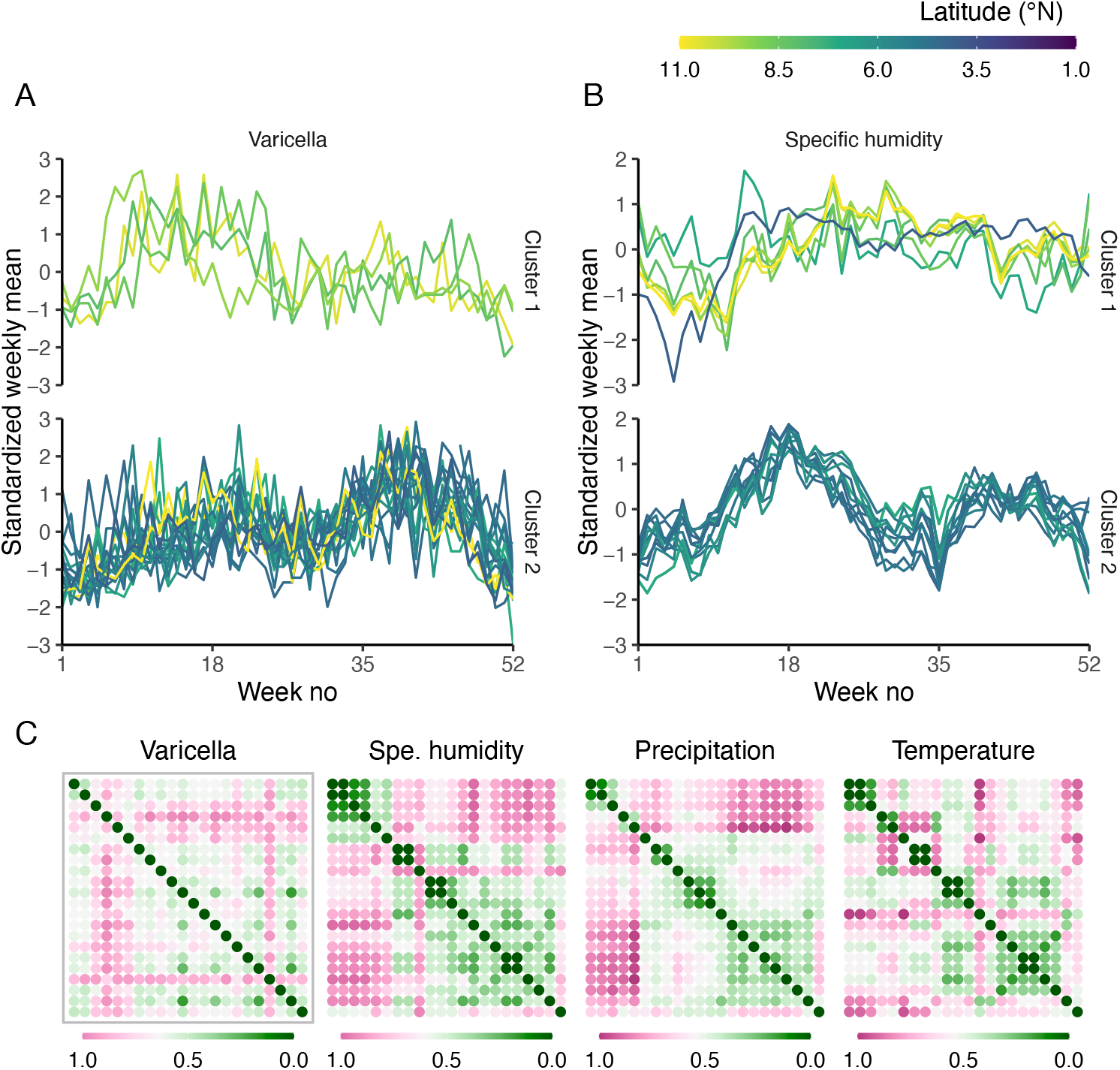
Correlation between spatial heterogeneity of varicella seasonality and that of climatic variables. Standardized weekly mean varicella reports (A) and specific humidity (B) independently clustered in two municipality groups. (C) Between-municipality similarity (based on Euclidean distance) matrices of varicella, specific humidity, precipitation, and temperature (from bottom to top and left to right, the municipalities are ordered by increasing latitude).

To strengthen the evidence for a link between climate and varicella seasonality, we ran a series of Mantel tests to assess the correlation between the spatial similarity matrix of varicella seasonality and that of every climate variable (Fig. 3A). We found evidence of strong positive correlations with all humidity variables and precipitation, but not with temperature (Table 1). We also ran additional analyses to test our assumption that distance between municipalities does not directly affect varicella seasonality (Fig. S5). Despite evidence of a positive correlation with distance from a direct Mantel test (Mantel statistic = 0·278, p-value = 0·017), this correlation became negligible after correcting for latitude or humidity in a partial Mantel test (Mantel statistic = 0·132 and −0·002, p-value = 0·131 and 0·456, respectively). Moreover, the correlation with humidity remained robust after controlling for latitude or distance (Mantel statistic = 0·316 and 0.337, p-value = 0·003 and 0·003, respectively). These results suggest that spatial variability in climate across municipalities, but not the spatial distance between municipalities, can explain the variation in the seasonality of varicella across Colombia.

**Table 1.** Correlation between the spatial heterogeneity of varicella seasonality and that of climate. The correlations were calculated using Mantel and partial Mantel tests on the dissimilarity matrices calculated using Euclidean distances between the time series of paired municipalities.

| Variable tested | Control variable | Mantel statistic | p-value |
| --- | --- | --- | --- |
| Specific humidity |  | 0.412 | 0.001 |
| Absolute humidity |  | 0.398 | 0.001 |
| Total precipitation |  | 0.319 | 0.008 |
| Relative humidity |  | 0.244 | 0.020 |
| Mean temperature |  | 0.077 | 0.225 |
| Specific humidity | Distance | 0.316 | 0.003 |
| Specific humidity | Latitude | 0.337 | 0.003 |
| Distance |  | 0.278 | 0.017 |
| Distance | Latitude | 0.132 | 0.131 |
| Distance | Specific humidity | -0.002 | 0.456 |

### A mechanistic model with transmission rate seasonally forced by humidity can reproduce the latitudinal gradient of varicella seasonality in Colombia

To further examine the impact of climate on varicella seasonality in Colombia, we formulated a simple mechanistic model in which the transmission rate of varicella infection was seasonally shaped by alternating school terms and school holidays (i.e., term-time forcing)^29^ and by variations of specific humidity (i.e., humidity forcing). In the absence of humidity forcing (Fig. 4A, left heatmap), the incidence of varicella gradually increased after the Christmas and mid-year vacations, resulting in a first peak around week 24 and a second, more pronounced peak around week 49 throughout Colombia. This scenario failed to recreate any latitudinal gradient. By contrast, a latitudinal gradient was predicted by simulations with a small effect of humidity forcing (where a 1-SD (standard deviation) increase in humidity resulted in a 4% decrease in the transmission rate of varicella infection, Fig. 4A, middle heatmap). Increasing the effect of humidity forcing resulted in a more definite latitudinal gradient, which broadly reproduced that observed in Colombia, with a contrast between southern and northern municipalities (Fig. 4A right panel and Fig. 4C and E) (where a 1-SD increase in humidity resulted in an 8% decrease in the transmission rate of varicella).

**Figure 4.**
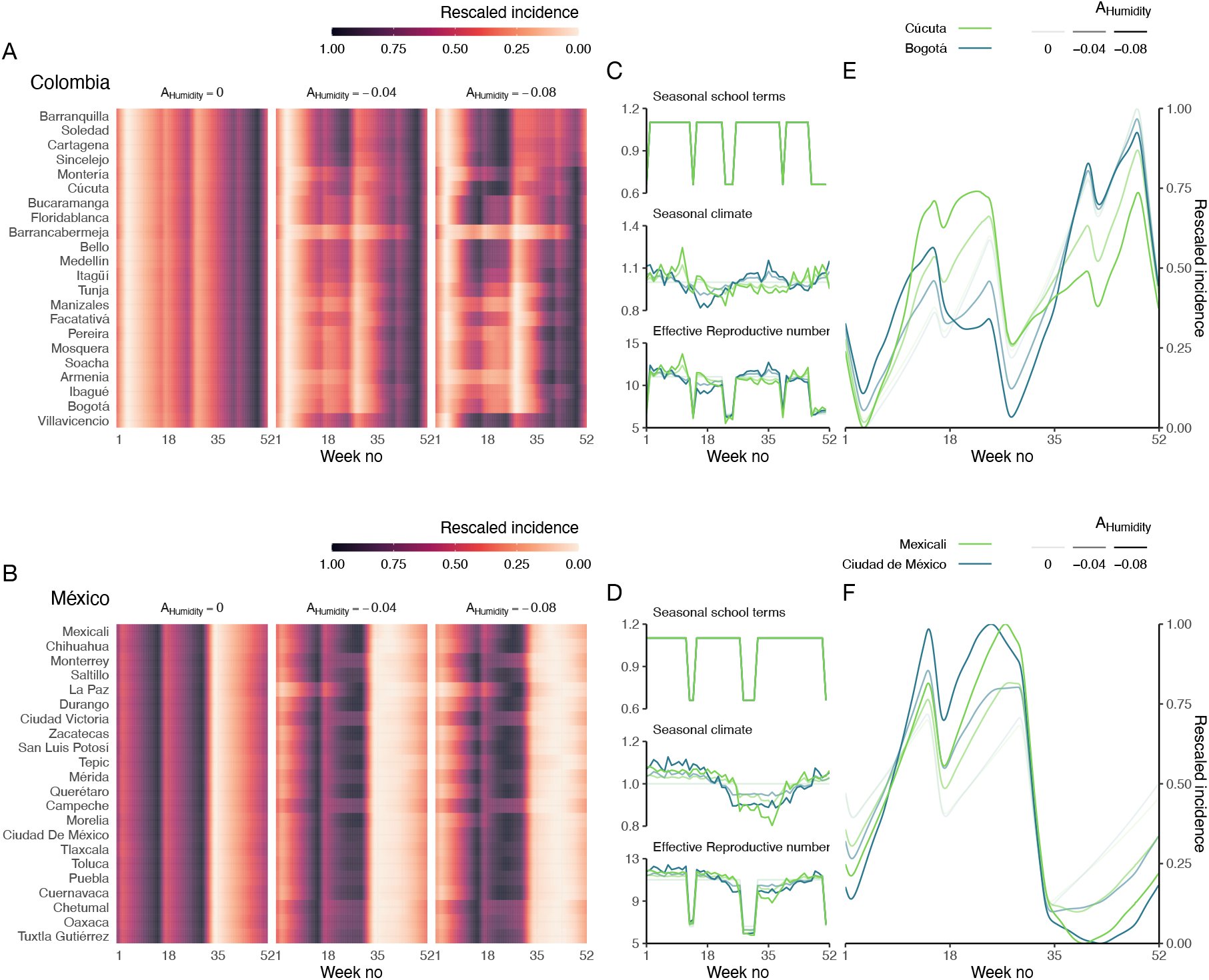
Transmission models predicting the impact of seasonal humidity and alternating school terms and holidays on the transmission rate can reproduce the latitudinal gradient of varicella seasonality in Colombia and México. Predicted varicella incidence (rescaled so that 0 is the minimum and 1 is the maximum number of cases per municipality) for all (A) municipalities in Colombia and (B) cities in México. (C and D) Estimated seasonal components of the transmission rate and (E and F) corresponding seasonal varicella profiles in the municipalities of Cúcuta (7·88° N latitude) and Bogotá (4·71° N latitude) and in the cities of Mexicali (32·6° N latitude) and Ciudad de México (19·4° N latitude). The municipalities are ordered according to their latitude. For all municipalities and cities, the model was run for a period of 200 years until equilibrium. The results displayed correspond to the last simulated year.

To complement our Colombian data, we simulated the dynamics of varicella in México, a sub-tropical country with variable climates but uniform subnational seasonal variations of humidity across 32 cities. In stark contrast to Colombia, the predicted seasonality of varicella did not vary across space, with a single peak around week 14 in all cities, as observed previously (Fig. 4 B, D and F, and Fig. S7).^30^ To further predict the importance of subnational heterogeneity of climates, we simulated the dynamics of varicella in all countries of Central America. In the northern countries of Belize, Guatemala, and El Salvador (located south of México), we predicted only small variations of varicella seasonality between the different cities. In the southern countries of Panamá, Costa Rica, Nicaragua, and Honduras, however, we predicted substantial subnational variability in varicella seasonality, echoing our findings in Colombia (Fig. S8). These results emphasize the need to pay careful attention to the subnational heterogeneity of climates when studying the seasonality of varicella.

## Discussion

Over the last decade, almost a billion children have had varicella worldwide—including 1 million in Colombia—and will harbor the virus lifelong in sensory neurons.^1^ Most associated hospitalizations and deaths occur in low- to middle-income countries. Many of them are located in tropical regions, which generally have limited surveillance data.^10^ Because of this data gap, research into the epidemiology of varicella in tropical regions is needed for appropriate health preparedness and resource planning.^1,31^ In this study, we examined the seasonality of varicella in the tropical climates of Colombia, based on an extensive dataset of weekly reports in children up to the age of ten in 25 municipalities during 2011– 2014. Using a series of descriptive statistical models, we showed that the seasonality of varicella incidence is markedly bimodal, with a latitudinal gradient in the amplitude of two peaks. We then used clustering methods and matrix correlation tests to compare the spatial variability in varicella seasonality to the spatial variability of five meteorological variables throughout Colombia. The estimated correlation was strongest for specific and absolute humidity, weaker for precipitation and relative humidity, and negligible for temperature. Finally, we showed that a simple dynamical model of varicella, where transmission was seasonally forced by specific humidity, could reproduce the observed seasonalities of varicella incidence across Colombia and México. These findings suggest that seasonal variations in humidity may explain the seasonality of varicella in Colombia and likely in other Central American countries. More generally, our results emphasize the need to carefully consider the subnational heterogeneity of climates when studying the seasonality of varicella.

We found evidence for a markedly bimodal seasonality of varicella, with a latitudinal gradient in peak magnitude correlated with spatiotemporal variations of humidity across the tropical climates of Colombia. This heterogeneity can be explained by the wide diversity of climates in Colombia, attributable to a unique combination of the Intertropical Convergence Zone (ITCZ), the Andes mountains, and the South American monsoon in this country.^32^ In contrast, we predicted that the spatially uniform humidity profiles of México resulted in rigidly unimodal varicella seasonality throughout the country’s subtropical climates. These predictions are confirmed by previous observations in México, including a modeling study.^30,33^ Together with our predictions across Central America, these results suggest that varicella seasonality is less spatially homogeneous in tropical countries of this region. A practical consequence is that the observed epidemiological dynamics of varicella may be blurred in nationwide reports, which have been used in previous studies to characterize varicella seasonality worldwide and to estimate the impact of the introduction of the varicella vaccine (2015) in Colombia.^4,34^ Our results, therefore, show the need to carefully consider the subnational heterogeneity of varicella for epidemiological studies and public health policy.

Beyond tropical regions, the seasonality of varicella varies with latitude globally, with a peak in temperate regions during March-May in the northern hemisphere and during October–December in the southern hemisphere.^4^ Although a single peak is observed in most temperate countries (like in European countries, the USA, Australia, and South Africa), two peaks are observed in other countries at similar latitudes (like the UK, Turkey, Japan, and China).^7,35–37^ Previous modeling studies have attributed these patterns to seasonal variations in host contacts (due to alternating school terms and school holidays),^38,39^ temperature,^7,11^ and less frequently to precipitation,^8^ and humidity.^30^ In temperate regions, the impact of humidity can be difficult to assess because it is highly correlated with temperature.^14^ In contrast, tropical climates offer a quasi-experimental setting where humidity, but not temperature, varies seasonally (Fig. S2). Hence, our findings add to the body of evidence on the impact of humidity on varicella transmission dynamics. Emphatically, although we found little correlation with temperature in our dataset, these results do not rule out that temperature is an important determinant of varicella epidemiology in other locations. They rather suggest that the contribution of individual climatic variables may vary with latitude. More generally, we propose that together with other elements affecting transmission, like vaccination and sociodemographic factors, seasonal variations in host contacts and climate can jointly explain the global seasonality of varicella.

Our findings suggest that humidity can impact the transmission of varicella and shape part of its seasonality. As evidenced for many other infectious diseases, climatic variables may impact pathogen transmission through multiple biological mechanisms which can affect the pathogen, the individual host, or the host population.^19,40^ At the pathogen level, experimental evidence has shown that humidity can determine the environmental stability of multiple respiratory viruses, such as influenza A, coronaviruses, adenoviruses, and rhinoviruses.^41^ Nevertheless, the humidity range optimal for viral stability has been found to vary markedly between different viral species (see table S1),^41^ highlighting the need for experimental studies on VZV. At the individual host level, changes in humidity may alter the host physiology, for example, the integrity of the respiratory mucosa, as demonstrated for influenza A in mice.^42^ Finally, at the host population level, variations of humidity and precipitation—in particular during rainy seasons—might modify the frequency of social contacts between individuals.^43,44,[preprint]45^ Hence, further studies will be necessary to test the biological mechanisms described above and to elucidate how humidity might affect varicella transmission.

Several limitations of our study are worth noting. First, the cases were not laboratory-confirmed and might, therefore, lack specificity because they include zoster, in addition to varicella cases. Nevertheless, we reduced this possible misclassification by focusing only on children up to ten, in whom zoster is extremely rare.^46^ Second, because of its observational design, further experimental studies are warranted to unveil potential causal effects. Even though we used a DAG to formalize our causal assumptions, we may have overlooked other explanations for the seasonality of varicella. Hence, further experimental studies in animal models and epidemiological studies in human populations will be useful to confirm our findings regarding the impact of humidity on varicella transmission. Third, in our transmission model, we assumed an exponential relationship between humidity and the transmission rate of varicella infection. Because this relationship is simple and easily interpretable, it has often been used to analyze the seasonality of respiratory pathogens.^47^ However, also other relationships, like power functions, are possible.^30^ Future work could estimate the form of this relationship, for example, by using recently developed statistical inference methods to confront transmission models to incidence data.^28^ Finally, because of our focus on Latin America, our results may not generalize to tropical regions in other parts of the world. In particular, it would be especially informative to expand our study to tropical regions of Africa or Asia, where varicella incidence is high as in Colombia, but where the underlying transmission dynamics may substantially differ.^1,48,49^

In conclusion, our results demonstrated substantial variability in varicella seasonality across the tropical climates of Colombia. They further suggested that seasonal variations of humidity could capture most of the spatiotemporal dynamics of the varicella incidence in this country. The predictable occurrence of varicella during specific times of the year is of importance for epidemic preparedness. In addition, careful consideration of seasonal heterogeneity may be crucial for unbiased estimation of the impact of varicella vaccination programs, which were recently rolled out in Colombia. Our study supports future research on the epidemiology of varicella in Colombia and other tropical countries.

## Data Availability

All data produced are available online at the web pages of the SIVIGILA, IDEAM, NARR, and CHIRPS. Furthermore, all the aggregated data (to the level of municipality-week) and code are stored in Edmond, the open research data repository of the Max Planck Society, to ensure the reproducibility of the results and will be made immediately available following publication.

http://portalsivigila.ins.gov.co/Paginas/Vigilancia-Rutinaria.aspx

http://www.ideam.gov.co/

https://www.ncei.noaa.gov/products/weather-climate-models/north-american-regional

https://www.chc.ucsb.edu/data/chirps

## Data sharing

All the data is freely available on the web pages of the SIVIGILA, IDEAM, NARR, and CHIRPS. Furthermore, all the aggregated data (to the level of municipality-week) and code are stored in Edmond, the open research data repository of the Max Planck Society, to ensure the reproducibility of the results and will be made immediately available following publication.

