## Supplements for "The seasonality of varicella in the tropical climates of Colombia: A statistical and mathematical modeling study"

---

Laura Andrea Barrero Guevara 1,2,*, Elizabeth Goult 1, Dayanne Rodriguez 3, Luis Jorge Hernandez 3, Benedikt Kaufer 4, Tobias Kurth 2, Matthieu Domenech de Cellès 1

1. Max Planck Institute for Infection Biology, Infectious Disease Epidemiology group, Charitéplatz 1, Campus Charité Mitte, 10117 Berlin, Germany
2. Institute of Public Health, Charité–Universitätsmedizin Berlin, Charitéplatz 1, 10117 Berlin, Germany.
3. Medicine Department, Universidad de los Andes, Bogotá 111711, Colombia
4. Institute of Virology, Freie Universität Berlin, Robert-von-Ostertag-Str. 7, 14163 Berlin, Germany

*Corresponding author: Laura Andrea Barrero Guevara, Max Planck Institute for Infection Biology, Charité-platz 1, Campus Charité Mitte, 10117 Berlin, Germany. E-mail address:

---

### S1 Supplementary data

#### S1.1. Varicella data

In Colombia, healthcare institutions report clinically confirmed varicella cases to the national surveillance system (*Sistema Nacional de Vigilancia en Salud Pública*, SIVIGILA), which in turn publishes online the freely available unidentifiable individual data.[1,2] The SIVIGILA works under the Colombian National Health Institute (NHI) in cooperation with the Pan American Health Organization (PAHO) and collects weekly data from public and private healthcare provider institutions. According to "Law 1712 of 2014 for Transparency and the Right to Access Public Information", the SIVIGILA database is available publicly online. The database provides 1) aggregated data, consisting of municipality-level, weekly time series of varicella cases, and 2) individual data without personal identification information from 2007 to date.[3,4] We aggregated the individual data by week and municipality, and all the analyses were performed using the aggregated data, which is available in the Edmond repository.

A varicella case is clinically defined by the SIVIGILA as an acute onset illness that initiates with moderate fever and small erythematous macules that evolve into papules, water-clear vesicles, yellowish pustules, and finally, crusts. The cases are evaluated by a healthcare professional and can be epidemiologically linked to another case. A varicella case is then stored under the 831 code in the SIVIGILA database.

#### S1.2. Spatial resolution and coordinates

The spatial resolution was defined at the level of the municipalities for Colombia, considering that the varicella data is reported at the same level. For México and Central America, the analyses were performed for the capital city of the first-level administrative divisions of each country. For Colombia, coordinates of the centroid for each municipality were obtained from the Colombian National Institute for Hydrology, Meteorology, and Environmental Studies (IDEAM) database.[5] For México and Central America, coordinates of the capital cities were obtained from Google Maps.

#### S1.3. School terms and school holidays data

We obtained the school information from 2014 for Colombia from the Education Ministry website.[6] Data for Mexico and other Central American countries (Panamá, Costa Rica, Nicaragua, Honduras, El Salvador, Guatemala, and Belize), were obtained likewise.[7–13]

---

### S2 Supplementary methods

#### S2.1. Transmission model formulation

We formulated a Susceptible-Exposed-Infected-Recovered (SEIR) model of varicella transmission, including school terms and climate as sources of seasonality for the transmission rate. The school terms driven by the alternation between school terms and school holidays were modeled using a square wave:

$$S_1(t) = \frac{1 + A_1 Term(t)}{1 - A_1(1 - 2p_{school})}$$

Where Term(t) equals 1 during school terms and –1 during school holidays. The parameter $A_1$ represents the amplitude of term-time forcing, fixed to 0.25. Climate forcing was modeled as:

$$S_2(t) = e^{A_2 Climate(t)}$$

where Climate(t) is the weekly standardized specific humidity, and $A_2$ represents the amplitude of the climate forcing, fixed to 0, –0.04 and –0.08.

The model is then represented using the following system of ordinary differential equations:

$$\begin{aligned}
\dot{S} &= \mu - (\lambda(t) + \mu)S \\
\dot{E_1} &= \lambda(t)S - (2\sigma + \mu)E_1 \\
\dot{E_2} &= 2\sigma E_1 - (2\sigma + \mu)E_2 \\
\dot{I_1} &= 2\sigma E_2 - (2\gamma + \mu)I_1 \\
\dot{I_2} &= 2\gamma I_1 - (2\gamma + \mu)I_2 \\
\dot{R} &= 2\gamma I_2 - \mu R
\end{aligned}$$

The seasonal force of infection is given by:

$$\lambda(t) = R_0 \gamma S_1(t) S_2(t)(I_1 + I_2)$$

For all simulations, the seasonal model was initialized at the values of the endemic equilibrium of the corresponding seasonally unforced model. The model was then simulated for 200 years, and the last year of the simulation was used for assessing the seasonality of weekly cases.

---

### S3 Supplementary figures

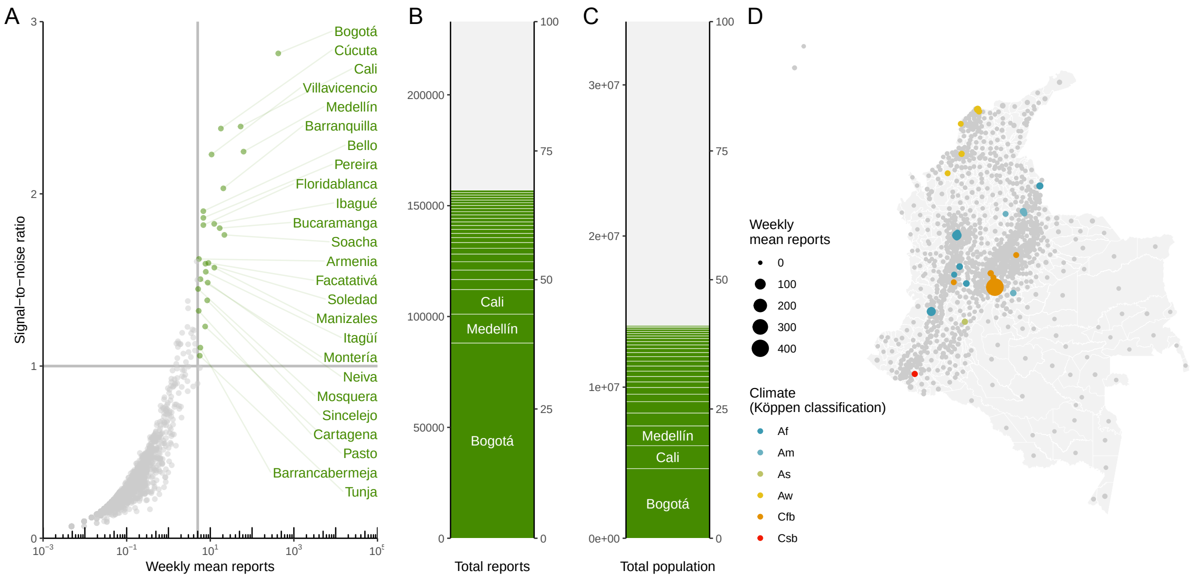

**Supplementary figure 1. Selection of municipalities according to varicella reports and signal-to-noise ratio.** For definiteness, we selected the municipalities with a signal-to-noise ratio (mean to standard deviation ratio of the weekly reports) over one and an average of at least five cases/week. (A) 25 municipalities met the criterion and collectively included (B) 67.4% of the total reports of varicella, (C) 41.2% of the total study population (children up to the age of ten). (D) Map of included municipalities, the 25 municipalities included six of the 17 Köppen–Geiger climatic classifications most commonly found in the tropics.

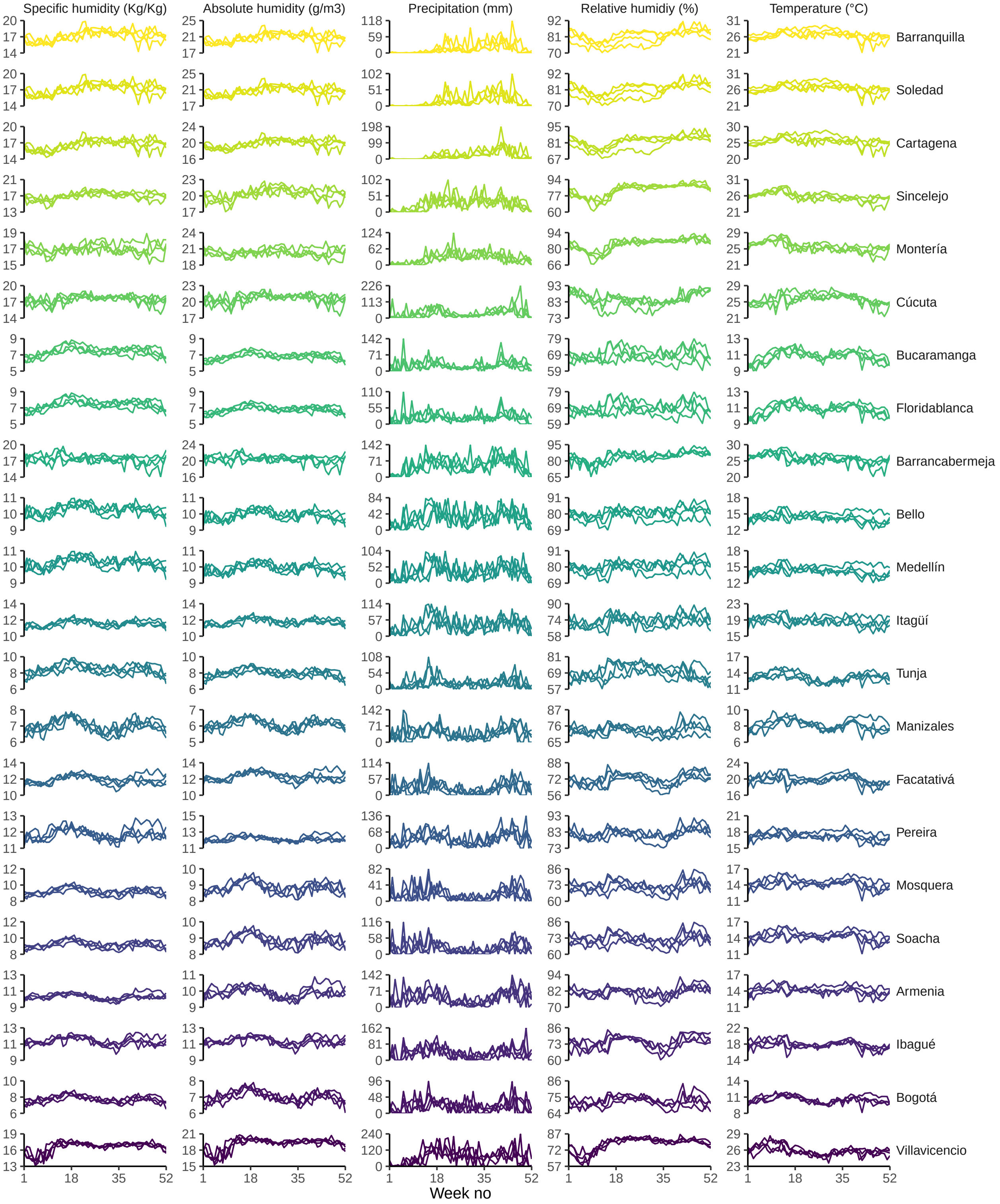

**Supplementary figure 2. The seasonal climate in municipalities of Colombia.** For the study period (2011–2014), we accessed the weekly specific humidity (expressed in g/kg), absolute humidity (g/m3), relative humidity (%), and temperature (°C) from the North America Regional Analysis (NARR) and the weekly precipitation (mm) from the Climate Hazards Group InfraRed Precipitation with Station data (CHIRPS).

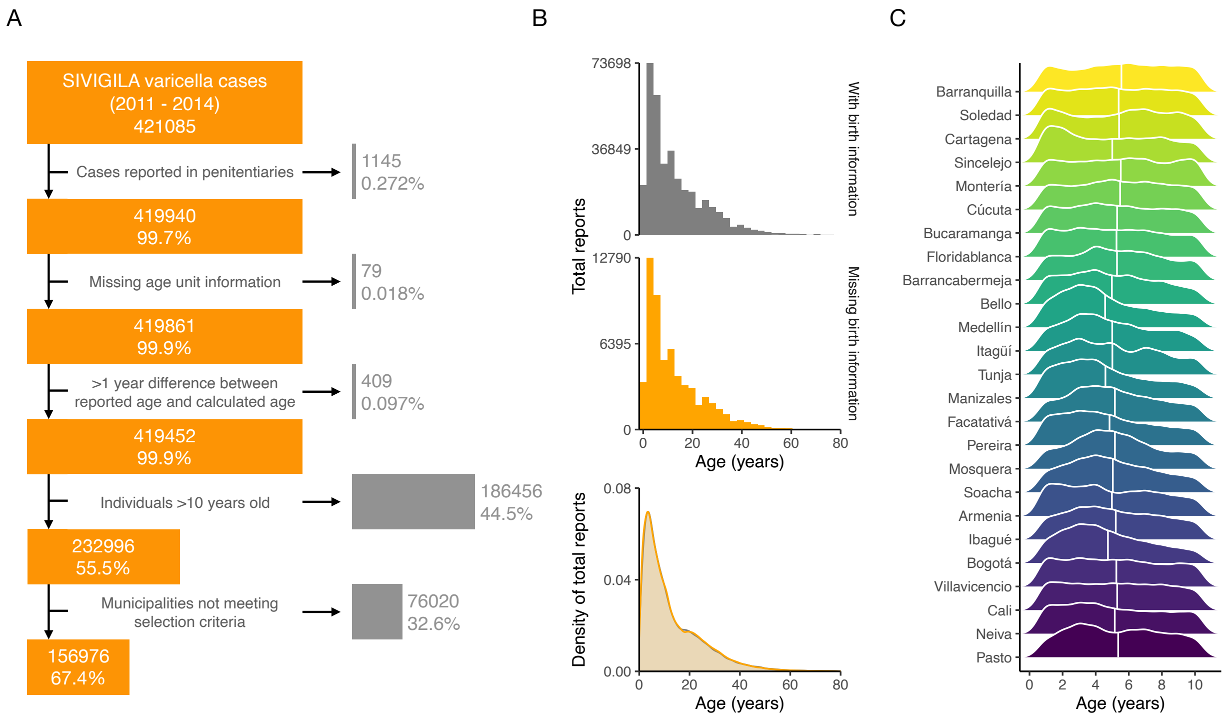

**Supplementary figure 3. Data flow chart on varicella reports.** We accessed varicella cases for the study period (2011-2014) from the Sistema Nacional de Vigilancia en Salud Pública (SIVIGILA). (A) Data flow chart. (B) For 60,257 cases that reported age but no birthdate information, we assumed that the age was not misclassified as their distribution was not different from the cases with birthdate data. (C) The age distribution of varicella infections across municipalities in Colombia in children up to the age of ten. The vertical white line represents the mean age at infection.

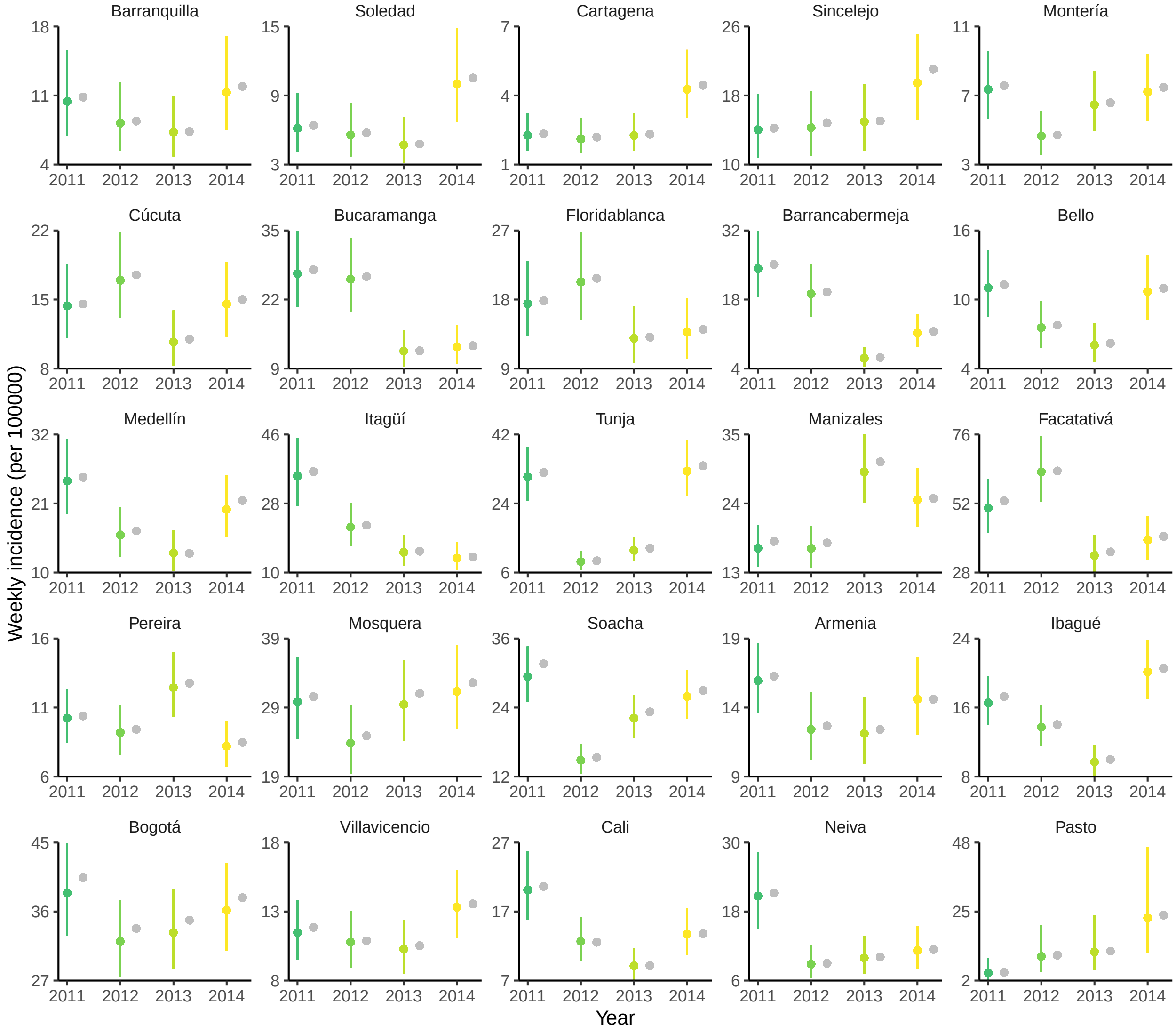

**Supplementary figure 4. Predicted weekly incidence per year in each municipality of Colombia.** The colored points (and colored lines) represent the point estimates (and 95% confidence intervals) of the average weekly incidence for every year and every municipality, estimated from the GAMs. The gray points indicate the observed average weekly incidence calculated from the data.

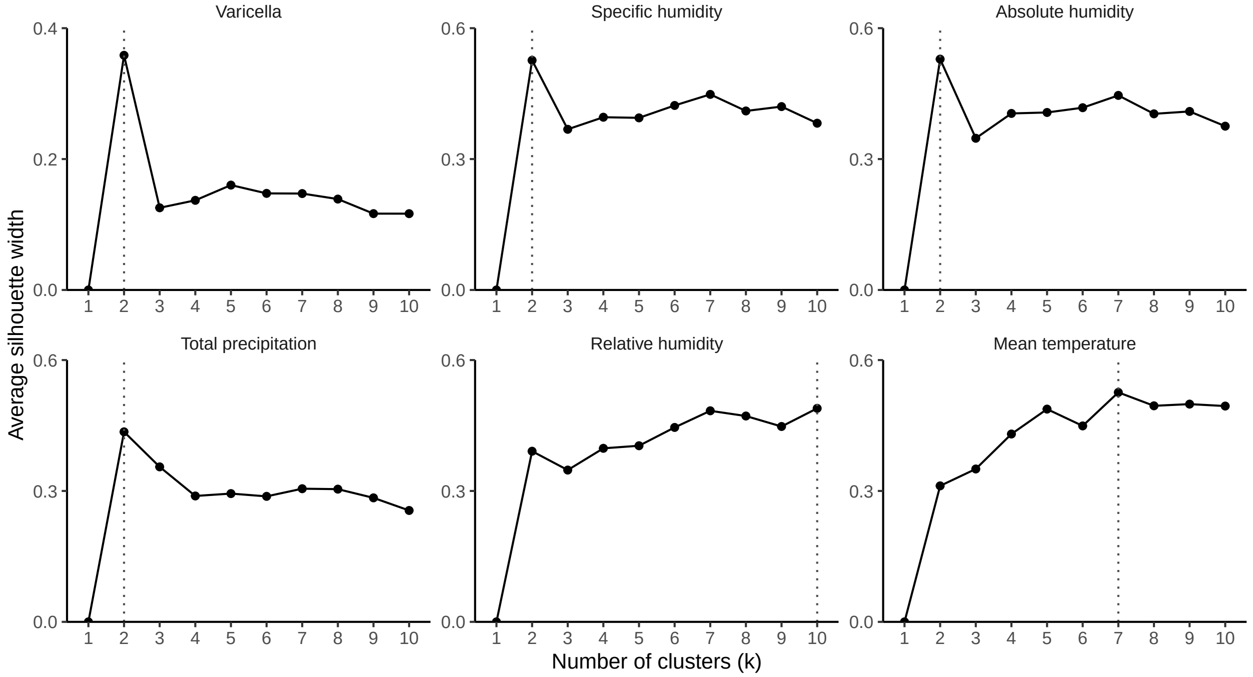

**Supplementary figure 5. The optimal number of clusters for varicella and each climatic variable.** Average silhouette widths were calculated for 1 to 10 clusters per variable. The dotted line shows the selected number of clusters.

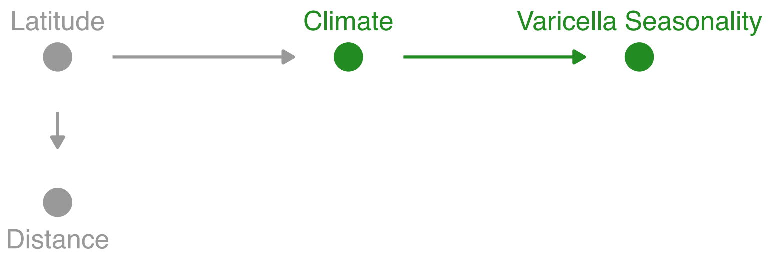

**Supplementary figure 6. Directed acyclic graph (DAG) on the effect of climate on the varicella seasonality.** Assumptions on the impact of climate on seasonality. Latitude and distance would have an indirect effect on seasonality.

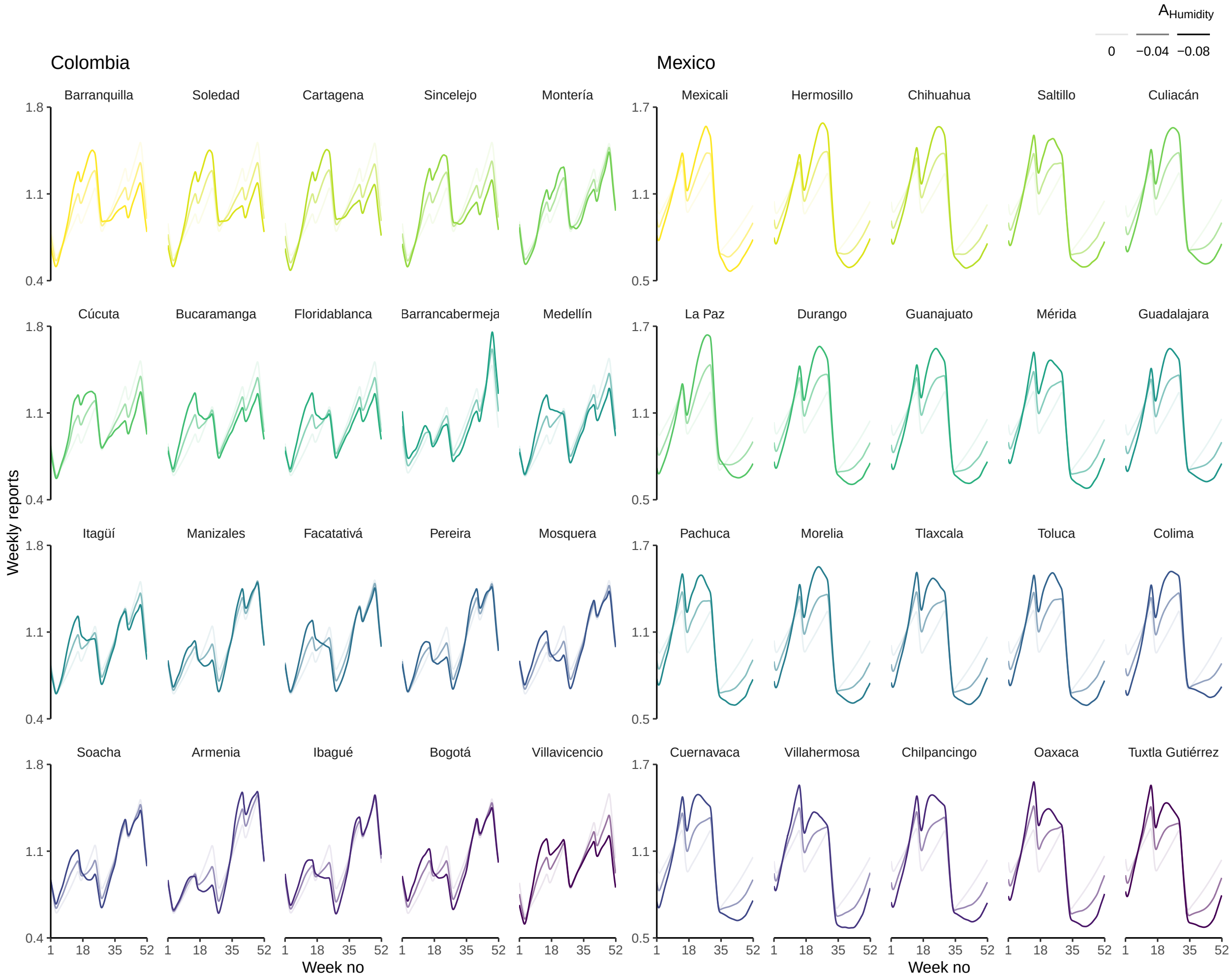

**Supplementary figure 7. Seasonal varicella profiles in the municipalities of Colombia and México.** Predicted varicella incidence produced using a transmission model including the impact of seasonal humidity and alternating school terms and holidays on the transmission rate of varicella.

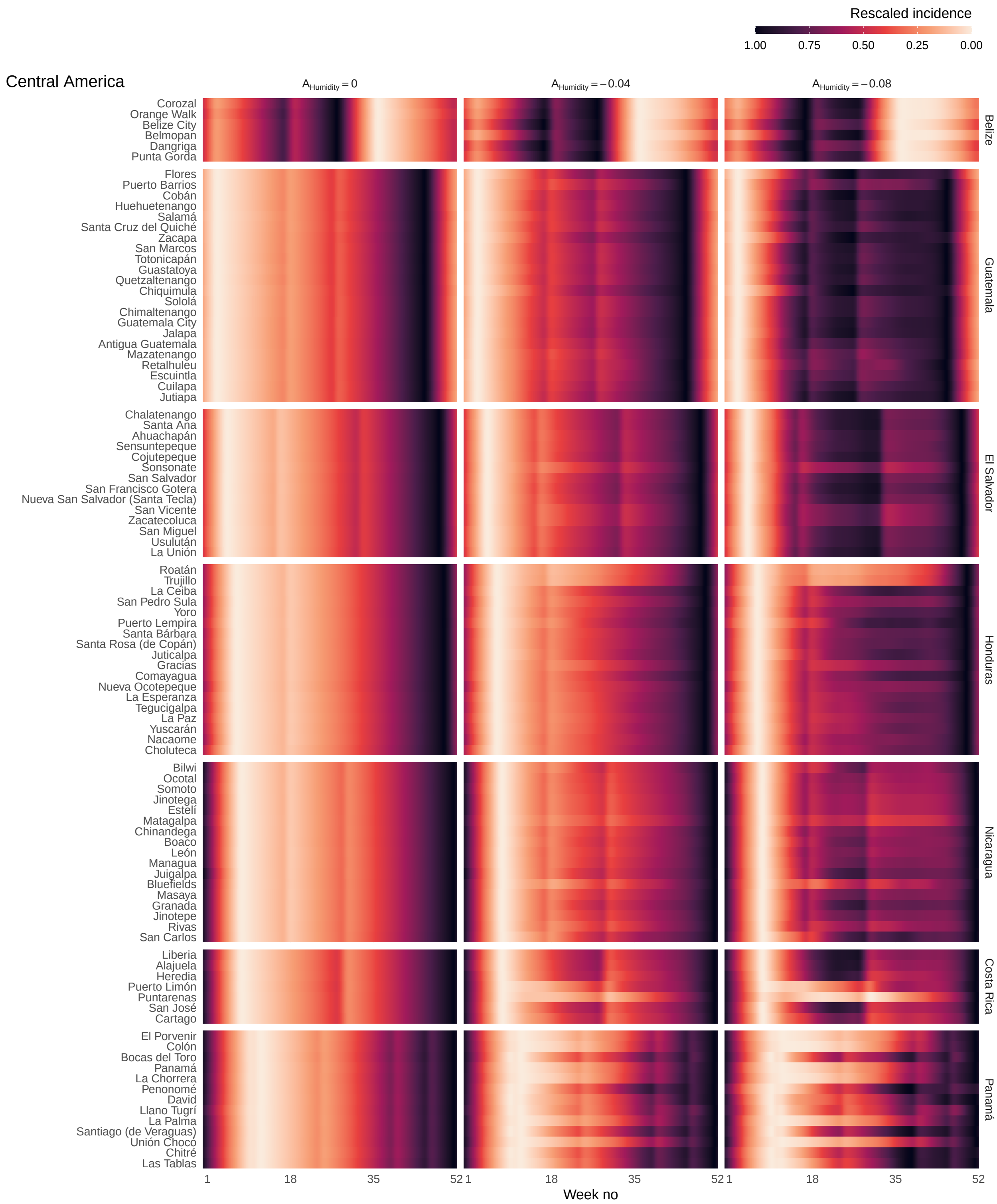

**Supplementary figure 8. Transmission models predicting the impact of seasonal humidity and alternating school terms and holidays on the transmission rate simulate the seasonality of varicella in countries of Central America.** Predicted varicella incidence (rescaled so that 0 is the minimum and 1 is the maximum number of cases per municipality) for all countries in Central America. The cities are ordered according to their latitude. For all cities, the model was run for a period of 200 years until equilibrium. The results displayed correspond to the last simulated year.
